# The Acceptability of a Tobacco Dependency Treatment for NHS Staff in the North East of England: A Mixed-Methods Study

**DOI:** 10.1101/2025.01.10.25320250

**Authors:** Caitlin Thompson, Kerry Brennan-Tovey, Caitlin Robinson, Rachel McIlvenna, Eileen FS Kaner, Sheena E Ramsay, Maria Raisa Jessica Aquino

## Abstract

**Aims:** High smoking rates and deprivation levels in North East of England led to an opportunity to pilot a tobacco dependency treatment offer for NHS Staff who smoke to make a supported quit attempt. The direct and indirect benefits to staff, patients, and NHS organisations are well documented. This study aimed to evaluate the acceptability of the service.

**Methods:** The service included up to 12 weeks of free Nicotine Replacement Therapy (NRT) and/or a refillable e-cigarette, motivational support, and premium access to the Smoke-Free App. Service evaluation used a mixed-methods design, combining the Theoretical Framework of Acceptability (TFA) questionnaire and semi-structured interviews with staff who had accessed the offer. Quantitative data were analysed using descriptive statistics and qualitative data via thematic analysis.

**Results:** 68 survey responses (*M_age_* = 43.92 years, *SD* = 12.27; Gender = 27.9% M and 72.1% F) reflected high acceptability (*M*= 4.59, *Mdn* = 5.00, *SD* = 0.72). Eighteen interviews (*M_age_* = 40.67 years, *SD* = 12.22; Gender = 38.9% M and 81.1% F) revealed four themes relating to service familiarity and ease of access, suitability of the NRT/e-liquid ordering service, the vape kit, and behavioural support.

**Conclusions:** The service was deemed highly acceptable, and service users’ experiences informed recommendations for improving future tobacco dependency services. This is the first known application of the TFA to an evaluation of a smoking cessation intervention and contributes to a broader body of research on reducing tobacco dependence.

## Background

The UK’s National Health Service (NHS) is the fifth largest employer in the world, with NHS England employing 1.5 million staff in February 2024^1^. While smoking rates amongst healthcare staff are typically lower than the general population, a substantial number of NHS staff are likely to smoke^1^. The Hiding in Plain Sight report^1^ highlighted that staff smoking costs the NHS approximately £206 million each year, comprising £101 million from sickness absence, up to £99 million from smoking breaks and £6 million in sickness treatment costs^1^. Providing support for staff smoking cessation could save the NHS on the costs of treating tobacco-related illnesses, which can also positively impact the productivity of staff^1^.

The North East North Cumbria (NENC) NHS Integrated Care Board (ICB) is one of the largest integrated care systems in the UK, employing 170,000 staff members. Across the NENC, staff smoking contributes to 36,710 hospital admissions and 14,288 premature deaths in the region annually^2^. This costs the NHS approximately £2.8 million in productivity, social, healthcare and fire costs across NENC ICB^2^. Addressing staff smoking remains a priority within the region, and doing so can help support the implementation of the NHS Long-Term Plan by promoting smoke-free environments for both staff and patients^3^.

In the NENC, smoking prevalence rates remain higher than the England average (13.0% vs 12.7%, respectively)^1^. The NENC has one of the highest deprivation levels in the country^2^. Although rates are trending downwards, figures continue to mask inequalities, with higher rates of smoking amongst those with lower socioeconomic status, and smoking continues to be the leading driver of health inequalities^4^. As such, smoking prevalence remains high amongst routine and manual workers, with 21.5% of routine and manual workers being smokers across the NENC Integrated Care Board (ICB)^5^. Supporting staff across all work settings to quit smoking remains a priority to address health inequalities in the region.

Limited access to quit-smoking support through work has been identified as a barrier to successful quit-smoking attempts^6^. Under national pilots funded by the NHS England, Greater Manchester Integrated Care Partnership launched a tobacco treatment offer for Greater Manchester-based NHS employees. The offer included free access to the Smoke-Free App and 12 weeks of nicotine replacement therapy (NRT) and/or a refillable vape device to staff^7^. Results demonstrated reduced smoking rates, measured as 12-week abstinence rates, by 37-39%^7^. The highest quit rates were reported for NRT/vape use in combination with the app, compared to the app alone^7^. Although promising, these findings do not lend insight into the acceptability of the service from the users’ perspective, including the barriers and enablers to engagement. Providing this evidence could help inform the sustainability of implementing such a service to staff working in NHS settings.

In light of NICE guidance^8^ and priorities outlined in the NHS Long Term Plan^3^, the NENC ICB implemented the NHS Staff Tobacco Dependency Offer (STDO) alongside the rollout of NHS England-funded tobacco dependency treatment services in acute inpatient, mental health inpatient, and maternity settings. The current evaluation aimed to address gaps in the evidence on the acceptability of tobacco dependency services for NHS staff by exploring the extent to which the NHS Staff Tobacco Dependency Offer (STDO) was acceptable for staff in the North East and North Cumbria (NENC) and identify any barriers/enablers to engagement which may affect successful quit rates.

## Methods

### The NHS Staff Tobacco Dependency Offer

NENC STDO was piloted from December 2021 to September 2023, with 1972 people taking up the offer across the NENC (supplementary materials). For context, at the time of the evaluation (September 2023), the NENC ICB employed 81,117 staff^9^. The service, however, also included staff employed by subsidiary companies, which may not be reflected in NHS workforce statistics. This is worth noting, given the portion of routine and manual workers within this subgroup of staff.

The offer provided NHS staff who smoke with up to 12 weeks of free NRT products and/or refillable e-cigarettes as well as access to free 12-week motivational support with a trained stop-smoking advisor. Access varied across the region; for example, some areas had specialist services, such as a team of advisors who saw smokers through a 12-week quit attempt within a Trust. In contrast, other areas commissioned services (i.e., pharmacies or GP practices) to deliver the intervention on their behalf. All staff were offered full premium access to the Smoke-Free app^10^ for nine months, which offers remote, flexible support 24/7.

### Design and Procedure

A mixed-methods evaluation was undertaken, consisting of quantitative cross-sectional survey work and qualitative semi-structured interviews. Participants were recruited by email and through the stop smoking service. Recruitment posters were shared through internal communications by members of the ICS NENC team. The survey took up to 15 minutes to complete (online and paper copies were available). At the end of the survey, participants were given an opportunity to leave a contact email address/phone number to participate in an interview. Participants were offered a £15 voucher following interview completion. Participants could also contact researchers to participate in an interview without completing the survey. Informed consent was taken from all participants. Data collection occurred between July to November 2023. Interviews were audio recorded and transcribed verbatim (by an external transcription company). Following the interviews, participants were provided with a debrief form.

### Participants

Convenience and snowball sampling techniques were used to recruit participants who met the following eligibility criteria: staff employed or a subsidy of one of the NENC NHS Trusts; self-identify as smoking tobacco products or as previously smoking tobacco products; currently engaging with the NENC STDO service or engaged but withdrew from the NENC STDO service or engaged and completed the course of treatment provided by the NENC STDO service. Recruitment was informed by information power^11^.

## Materials

### Survey

Acceptability was measured using a survey instrument based on the Theoretical Framework of Acceptability (TFA)^12^. The TFA questionnaire is a validated, brief and adaptable tool to measure intervention acceptability across a range of healthcare settings^13^. The TFA consists of eight constructs: Affective Attitude, Burden, Ethicality, Effectiveness, Coherence, Self-Efficacy, Opportunity Costs, and General Acceptability. Participants respond to the survey on a Likert scale from 1-5 depending on the context of the question, e.g., strongly disagree to strongly agree^12^.

### Interviews

Semi-structured, one-to-one qualitative interviews were used to gather experiences and perspectives of the service users who had engaged with the NENC STDO. Interviews followed a bespoke topic guide informed by the TFA^12^. Interviews lasted approximately 30-45 minutes and took place over Microsoft Teams or telephone, depending on participant preference.

## Data Analysis

### Survey

Data was managed using IBM SPSS Statistics (Version 27). Questionnaires were analysed descriptively, using absolute and relative frequencies. Descriptive Statistics (mean (*M*) and median (*Mdn*) along with standard deviation (*SD*), standard error (*SE*), and 95% confidence intervals for each of the seven TFA constructs were obtained. Higher scores on TFA constructs are suggestive of higher acceptability^13^.Three of the seven constructs were reverse scored (Burden, Ethicality, and Costs).

### Interviews

Interview data were managed using NVivo^14^, and were analysed using Thematic Analysis^15^. Deductive coding built upon the seven TFA constructs alongside inductive coding (i.e., coding from the data) to identify key themes relating to acceptability and user experience that might not otherwise fit into the TFA constructs. The authors recognise that their own experiences and perspectives may have influenced theme development, and to ensure rigour, two authors (CT and KBT) independently coded 10% of the transcripts. They agreed upon a shared codebook, which CT used to code the remaining transcripts.

### Ethics

Ethical approval for the study was gained from Newcastle University’s ethics committee (reference: 31229/2022).

## Findings

### Survey

133 responses (110 online; 23 paper) were received. Removal of duplicates, incomplete, or invalid responses (*N*=65), resulted in a final sample of *N*= 68. Of the 68, 64 respondents provided their age (*M* = 43.92, *Mdn* = 43.50, *SD* = 12.27). Table 1 provides an overview of participant characteristics.

**Table 1:**
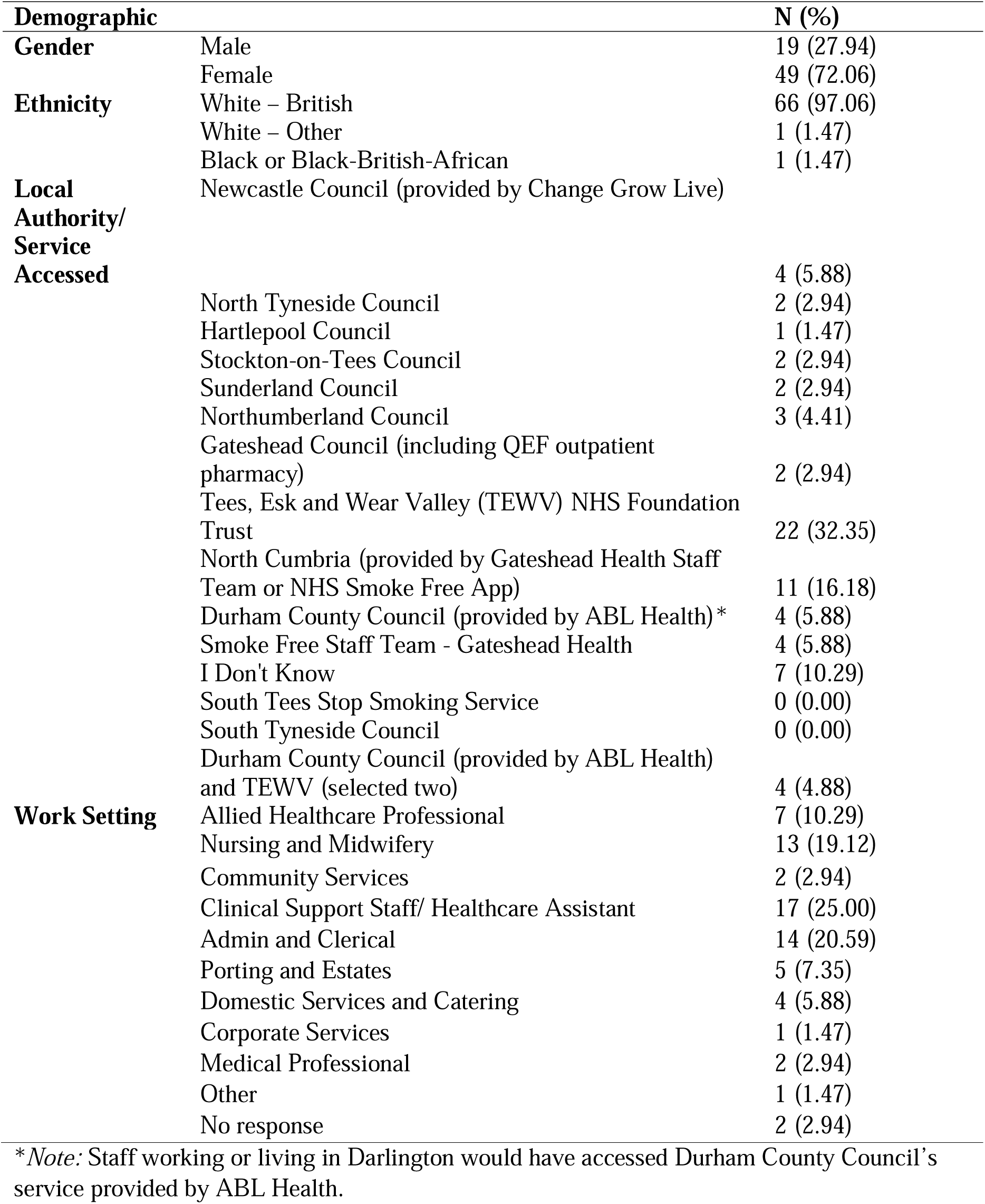
An Overview of Survey Sample Demographics (N= 68).

Survey results revealed that participants reported feeling ‘comfortable’ to ‘very comfortable’ in engaging with the STDO (Affective Attitude, see Table 2), (*M* = 3.877, *SD* = 1.552, *Mdn* =5.00). Participants also found it easy to engage with the intervention (Burden) (*M* = 4.108, *SD* = 1.331, *Mdn* = 5.00), and participants, on average, had ‘no opinion’ on whether the STDO had “ethical or moral consequences” (Ethicality) (*M* = 3.123, *SD* = 1.390, *Mdn* = 3.00).

**Table 2.**
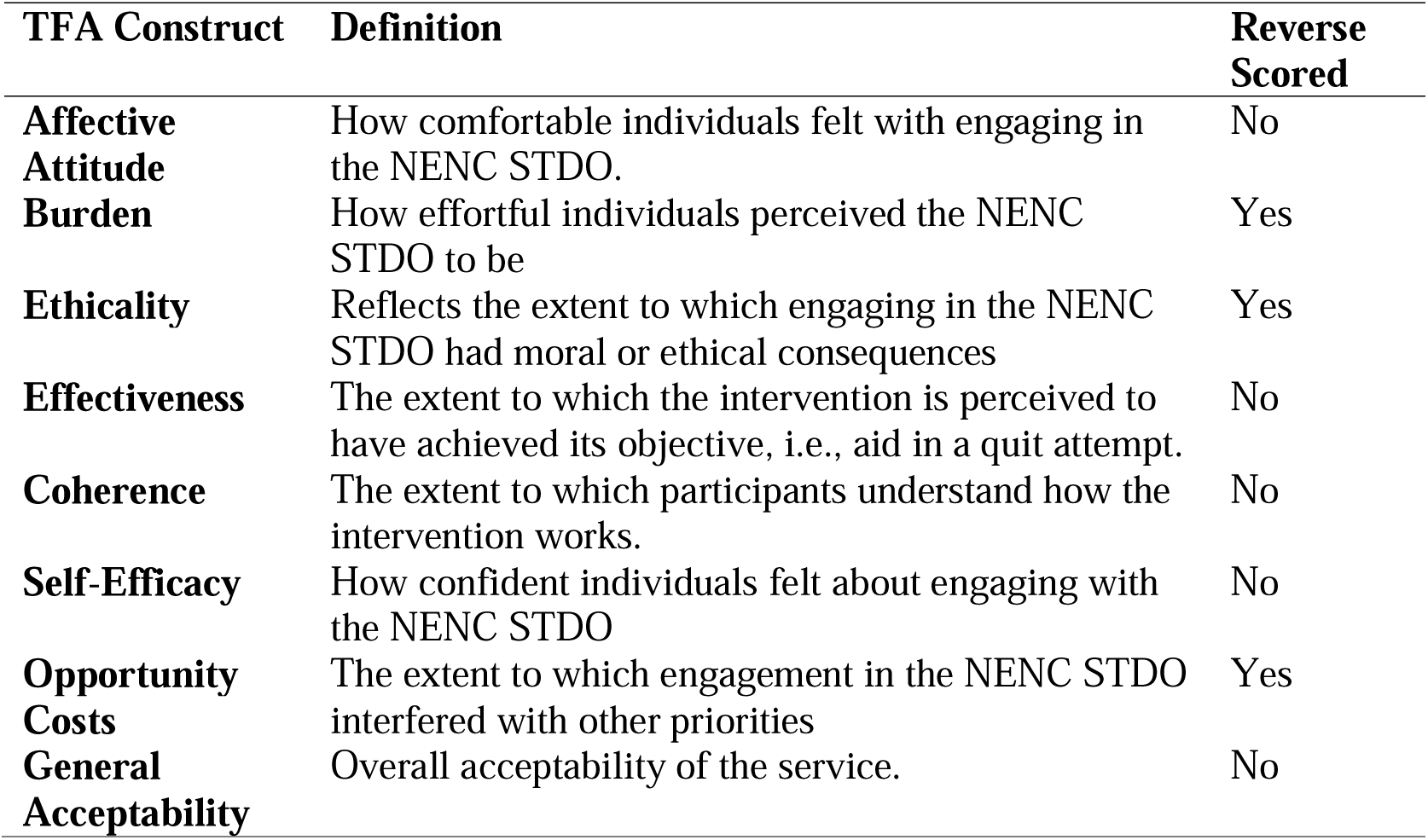
TFA Constructs, definitions and whether the construct is reverse scored.

Furthermore, participants tended to ‘agree’ that the service aided a quit attempt (Effectiveness) (*M* = 4.462, *SD* = 0.801, *Mdn* = 5.00). Staff also reported having a strong understanding of the package of treatment and support offered by the service (Coherence) (*M* = 4.477, *SD* = 0.783, *Mdn* = 5.00), and generally felt confident engaging with the service (Self-Efficacy) (*M* = 4.477, *SD* = 0.763, *Mdn* = 5.00). Responses also indicated that the NENC STDO did not interfere with existing priorities (Opportunity Costs) *M* = 3.754, *SD* = 1.480, *Mdn* = 4.00).

Finally, the survey revealed high overall acceptability (*M* = 4.585 (*SD* = 0.715, *Mdn* = 5.00) for general acceptability of the service. For an overview of descriptive statistics for each construct, see Table 3.

**Table 3.** Summary of Descriptive Measures of Each TFA Construct from Survey Results.

| **TFA Construct** | **Mean (SD)** | **Median** | **Standard Error** | **95% CI** |
| --- | --- | --- | --- | --- |
| **Affective Attitude** | 3.877 (1.552) | 5.00 | 0.196 | 3.486-4.268 |
| **Burden** | 4.108 (1.331) | 5.00 | 0.161 | 3.785-4.430 |
| **Ethicality** | 3.123 (1.390) | 3.00 | 0.172 | 2.780-3.467 |
| **Effectiveness** | 4.462 (0.801) | 5.00 | 0.101 | 4.260-4.663 |
| **Coherence** | 4.477 (0.783) | 5.00 | 0.082 | 4.312-4.641 |
| **Self-Efficacy** | 4.477 (0.763) | 5.00 | 0.096 | 4.285-4.668 |
| **Opportunity Costs** | 3.754 (1.480) | 4.00 | 0.185 | 3.385-4.123 |
| **General Acceptability** | 4.585 (0.715) | 5.00 | 0.090 | 4.405-4.765 |

### Interviews

A total of 18 interviews were conducted. Participants’ ages ranged from 23 to 63 years (*M* = 40.67, *Mdn* =37.5, *SD* = 12.22). Table 4 details interviewee demographic characteristics.

**Table 4.**
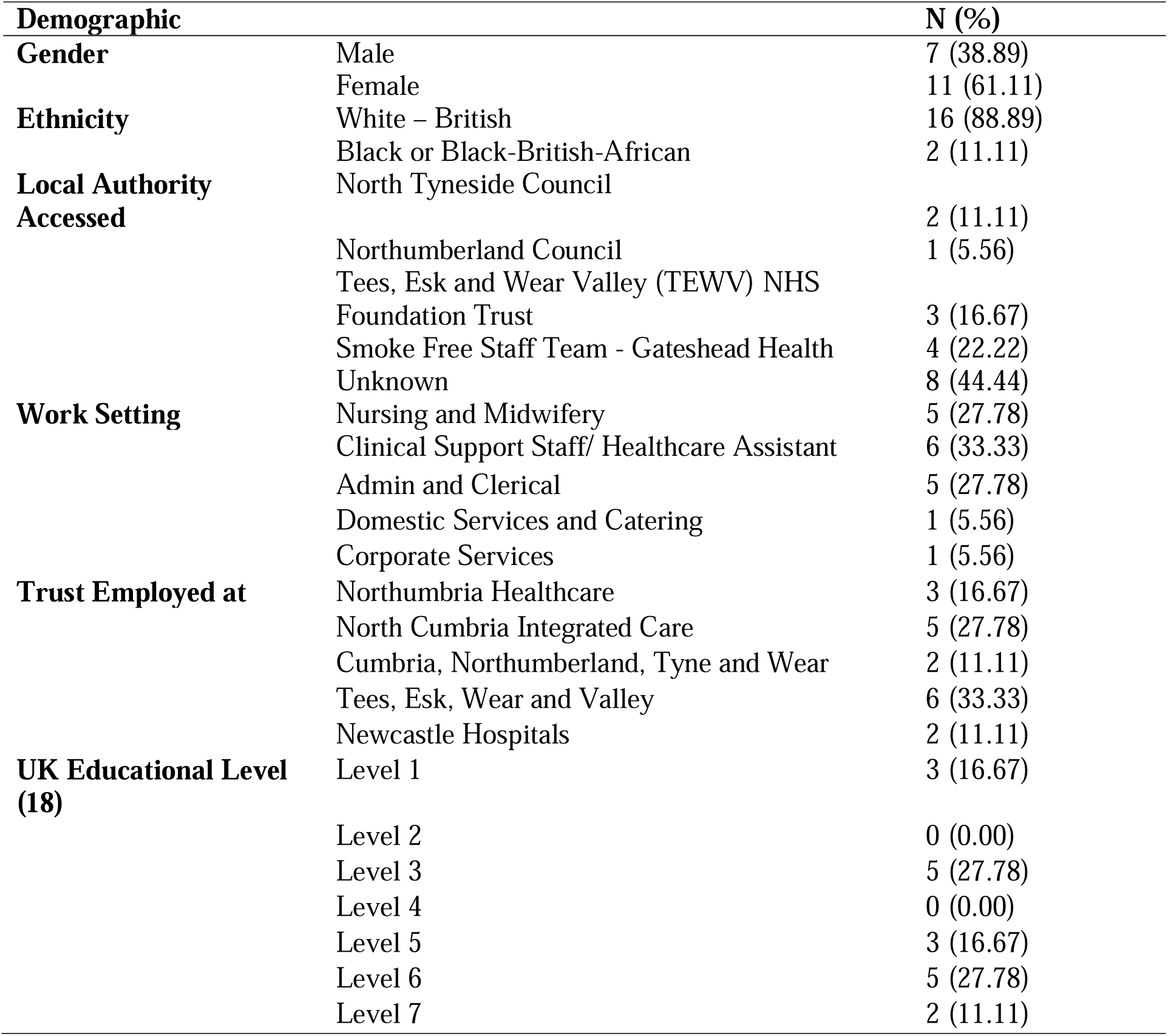
Interview Sample Demographics (N= 18).

### Themes from qualitative study

Four themes were identified relating to familiarity and ease of service access, suitability of the NRT/ E-liquid ordering service, suitability of the vape kit E-Liquid product, and suitability of the behavioural support offered by smoking cessation advisors. Each of the themes are discussed in turn below, with illustrative quotes provided. Quotes are presented with participant numbers, gender (M or F) and whether staff worked in clinical or non-clinical roles to uphold anonymity.

#### Familiarity with the service and ease of access

Some participants noted the service was well-advertised, whilst others reported that more staff needed to be made aware of the service depending on whether they were office-based or patient-facing. For example, participants noted that the service was advertised well for computer-based access, e.g., using QR codes, emails, and online staff pages, which could limit the type of workers it reached. However, once participants were acquainted with the service, accessing was thought to be easy, particularly for those with dedicated support on-site. Staff also reported convenience as they could access it through work, which motivated engagement:

> “No. It’s on our intranet… but obviously some people who we work with, say domestics and stuff like that, don’t really access the computers that often, and they might not know… I think sometimes, as well, having posters up and stuff like that… where people who haven’t got as much IT skills can still see it” (P005, F, Clinical);

> “Yeah, so it was very easy to do. Because I think they’re here from two o’clock on a Thursday until four o’clock so you can just pop down whenever you’re free” (P008, F, Non-Clinical).

#### Suitability of the NRT/e-liquid ordering service

Participants reported general satisfaction with NRT and E-Liquid options e.g., the amount offered from initial point of access, and throughout the duration of the 12 weeks. Participants were positive about being able to access their desired form of NRT. Furthermore, participants reported general acceptability and speed of the service’s ordering and delivery service for E-Liquids.

> “There was quite a good choice of liquids really…any flavour that you wanted in your milligrams of nicotine that you wanted” (P015, M, Clinical);

> “So I’ve got in touch, and it was really efficient, I filled out some paperwork and that, and then I think it was within two days I got a vape that came through with a month’s liquid, which was great” (P005, F, Clinical).

#### Suitability of the vape kit offered

Participants who accessed the e-cigarette reflected on the quality of the vape kit offered. For example, some experienced technical issues associated with their use. Many, however, complimented the vape kit as providing a promising starting point for their quit journey, with others reporting the vape kit as promoting a continued quit beyond the service. The free vape kit was also seen as a motivating factor for beginning their quit journey.

> “…the first vape that I got given wouldn’t charge so initially it was fine but then it wouldn’t charge… I ended up having to ring and they gave me some advice… And that got that going again and then I replaced it a third time. In those three months, I replaced the vape three times.” (P013, F, Clinical).

> “I think it’s a good service and as I say I think for me I obviously got the kind of free equipment and juice to get started but from that, that’s just been the kind of, the start really of kind of kickstarting me into stopping and it has been successful for me”. (P003, M, Non-Clinical).

#### Suitability of the behavioural support offered (smoking cessation advisors and app)

Many participants described good communication with Smoking Cessation Advisors, commenting that the regular contact helped to facilitate a successful quit. Some participants suggested that they could have benefitted from a stronger rapport being built, possibly with advisors having greater knowledge and experience (e.g., the effects of vaping). Also, some participants reported having wanted further behavioural support from the service.

There were mixed views regarding modalities of accessing the service. Participants found telephone contact with Smoking Cessation Advisors less acceptable than other modalities e.g., face to face. However, some reported accessing in-person drop-in Smoking Cessation Advisors appointments as challenging (e.g., preferring a private location for appointments). Participants suggested methods of improving accessibility, for example, signposting to peer support available, e.g., through the Smoke-Free app or creating peer networks to connect with other people accessing the service.

Concerning the Smoke-Free app, some participants reported being able to utilise tips offered on the app to aid their quit, whilst other comments suggested that it may not be suitable for supporting a quit attempt alone, working best in conjunction with other support, e.g., behavioural and/ or NRT.

> “I think that I anticipated more. I mean obviously you’re not going to do too much because people are busy but I thought there would have been more… even at nearly 40 you still like a little bit of praise every now and then” (P003, M, Non-Clinical);

> “Like a cessation support group, with other smokers going through, we can go and talk about their experiences…Then from a support group people create like WhatsApp chats or teams chats or whatever…” (P001, F, Clinical)

## Discussion

Survey findings revealed that overall, participants found the service to be acceptable. Qualitative findings reflect potential barriers to service engagement and provide insight into service user experience of the product and support offered. Barriers such as service advertisement were raised, with patient-facing staff possibly benefiting more from word-of-mouth/poster advertisement. Furthermore, the quality of the vape kit was reported on, alongside the importance of solid rapport building with advisors. Enablers of the service, such as ease of NRT ordering, access to a range of choices, and the fact that service was free and available through work, were reported as key strengths.

Previous studies have shown that smoking cessation services, which offer a combination of vape/NRT and behavioural support from an app, can help motivate successful quits and facilitate the move towards a smoke-free NHS workforce^6^, in line with the goals set out by the NHS Long-Term Plan^3^. Furthermore, a pharmacy-supported e-cigarette programme in the North West found that service users greatly appreciated the free device given, with financial savings playing a pivotal role in motivation to engage in a quit-smoking attempt^16^. Similarly, both a scoping review, a systematic review, and a meta-analysis of workforce tobacco dependency services found that workplace interventions are effective methods of motivating a quit-smoking attempt, with both recognising financial costs as key barriers to implementation^17,18^.

Previous studies have also demonstrated that strong motivational counselling has been found to improve long-term goal attainment in workplace stop-smoking services^18^. Having non-stigmatizing, positive behavioural support has also been found to reduce potential feelings of shame/stigma felt by staff seeking support from embedded workplace stop-cessation services^19^.

It is understood, therefore, that free, accessible services which offer a combination of NRT/e-cigarette and behavioural support can successfully motivate quit-smoking attempts. Overcoming barriers to obtaining stop-smoking support, such as embedding stop-smoking services in the workplace, as recommended by NICE guidelines^8^, is also evidenced as effective for service adherence. Less, however, is understood about the acceptability of embedded stop-smoking services within NHS workplaces. This study, therefore, adds to current understanding by providing the first known application of the TFA to evaluate the acceptability of an intervention package for smoking cessation embedded within NHS work settings.

Findings from this evaluation can be used to tailor future smoking cessation services for healthcare staff more generally. For example, findings highlight the need for services to be widely promoted using a combination of word-of-mouth, posters, and email/internet-based advertisement, with consideration for patient-facing workers with limited email/computer access. This is particularly important given that smoking patterns are highly socioeconomically patterned, with those working in non-clinical settings i.e., routine and manual workers demonstrating higher smoking rates^1^. Further work on service uptake, e.g., staff profession, to fully assess the reach of the service, is required, which was beyond the scope of the current evaluation.

In support of the findings from Greater Manchester^6^, qualitative accounts suggested that the Smoke-Free app is insufficient to support quitting smoking alone; rather, it is best used in conjunction with NRT and/or behavioural support. This is in line with current NICE guidelines, which recommend a combination of NRT and behavioural support (individual and group) in stop-smoking cessation services for an increased likelihood of positive clinical outcomes^8^. Interview findings also revealed a need for strong communication and rapport with advisors providing behavioural support. Participants additionally provided recommendations to strengthen behavioural support offered by the service, for example, through establishing network connections with other service users, to provide and receive peer support. Participants also noted different preferences for contacting Smoking Cessation Advisors, i.e., face to face/telephone, which could have facilitated rapport.

The following practice and policy recommendations for improving the acceptability of future stop-smoking services for healthcare staff have been developed, which could be applicable to wider smoking cessation services: Service promotion should be made widely accessible across different settings and include a variety of formats; To maintain service user engagement, formal and informal support should be available (e.g., on-site advisors, peer networks, bookable appointments); To support service delivery, Smoking Cessation Advisors should have regular training to ensure knowledge and information given is based on contemporary evidence; Finally, to maintain service quality, opportunities for providing feedback should be made available and used to inform service improvement.

### Limitations of this study

The study has some limitations. Firstly, there were challenges in recruiting participants for both the survey and the interviews due to the timing of the evaluation, i.e., there was a reduced number of staff accessing the service at the point of evaluation, leading to a smaller pool of potential participants. Challenges were overcome using different recruitment channels (i.e., email, posters, and word-of-mouth) and working closely with key contacts within services to recruit participants. It is also worth noting the drop-out rate from the survey sample, as although 133 survey responses were received, only 65 eligible responses were included in the analysis, accounting for incomplete responses and/or duplicates. The drop-out rate may reflect the feasibility of survey completion. It may also reflect the views of a limited pool of participants, i.e., those motivated to complete or those who had a positive experience of the service.

Furthermore, using the TFA presented challenges, for example, applying constructs to the evaluated service (e.g., the Ethicality construct). For example, the original TFA survey was trialled on the delivery of a COVID-19 vaccine^13^. The ethicality construct, i.e., an intervention’s fit with an individual’s values^12^, may be limited due to the moral and ethical differences and consequences of vaccination acceptability compared to that of a smoking cessation service. Future research is recommended to improve the application of TFA constructs for smoking cessation and other similar interventions to ensure accessible and applicable wording of survey questions. Lastly, it was beyond the study’s remit to validate the service outcomes, i.e., using quit data either as self-reported quit or as validated quit (CO monitoring).

### Conclusion

The evaluation demonstrates that the NENC STDO was deemed acceptable by NHS staff. Service users found the NRT/vape products and behavioural support to be accessible, with many service users wanting the service sustained, possibly for extended periods, with greater amounts of behavioural support being offered. Further research on services across different healthcare settings, and potentially further analysis of service uptake across work professions, is needed. The current study provides the first known application of the validated TFA framework to a smoking cessation intervention.

## Supporting information

Supplementary Materials 1 - Interview Topic Guide

Supplementary Materials 2 - Survey

## Data Availability

Anonymous survey data (as an SPSS file) will be shared on reasonable request to the corresponding author. Interview data cannot be shared publicly due to transcripts containing potentially identifying material and, therefore, cannot be shared to protect the anonymity and privacy of participants.

