## Supplementary Materials 1 - Interview Topic Guide for "The Acceptability of a Tobacco Dependency Treatment for NHS Staff in the North East of England: A Mixed-Methods Study"

### Evaluating the North East and North Cumbria NHS Staff Tobacco Dependency Offer

Thank you for agreeing to take part in this interview, and for giving up your time. We are inviting smokers who have recently accessed the NHS staff tobacco dependency offer, so I am pleased that we are meeting today. We are interested in finding out more about the stop smoking services you were offered or have previously accessed.

**History, Setting and Background (These questions are for all smokers)**

These first questions are going to be around you and the smoking services you have accessed before.

Are you currently a smoker? If so how long have you smoked? If you have recently quit, how long had you smoked?

Have you ever accessed any stop smoking services previously?

What help/support were you offered around smoking?

Did this include any advice on stop smoking services in your area, their contact details etc?

#### A) QUESTIONS FOR THOSE WHO SELF REFERRED INTO THE NHS-FUNDED TOBACCO DEPENDENCE STAFF OFFER SERVICE

Can you tell me how was the intervention explained to you?

What influenced your decision to commit to the intervention?

Can you tell me how the service ran for you?

Is there anything that you would have rather been offered?

How long were you interacting with this service?

Can you tell me about the services you are currently accessing now (to support you in your quit attempt)?

**Affective Attitude**: *How an individual feels about the intervention*

Can you tell me what you think/what your thoughts are on the service?

Can you tell me how you were feeling about starting the intervention?

Can you tell me how successful you found the service?

What are your thoughts in the need for this service to help people stop smoking?

**Burden**: *the perceived amount of effort required to participate in the intervention*

Do you think the service is easily accessible to staff?

Can you explain to me how much effort you thought you would need to engage with the service?

Can you explain to me how much effort was required on your part to engage with the service?

Where these the same?

**Ethicality**: *the extent to which the intervention has a good fit with an individual's value system*

Can you explain to me if you believe the service can work to help staff stop smoking?

Do you think the service is worth the effort required?

Why do you think that?

**Intervention Coherence**: *the extent to which the participant understands the intervention and how it works*

Are you able to explain to me the service that was offered to you?

Can you explain to me what was required of you to actively participate?

Can you tell me how the service was explained to you?

How well do you know the service now?

**Opportunity costs**: *the extent to which benefits, profits or values must be given up to engage in the intervention*

How did you find out about the service? Do you feel the service is easily accessible to others within the NHS?

Can you tell me what you had to give up to participate in the service effectively? (i.e., travel costs, time, friendships with smokers, work commitments)

Can you tell me about the additional 'costs' – not just financial, but also social and emotional, that you had to 'pay' to participate in the service?

Do you think the costs acquired were worth the outcome of being smoke free?

**Perceived effectiveness:** *the extent to which the intervention is perceived as likely to achieve its purpose*

Do you feel the intervention will be successful in helping you stop smoking?

Can you tell me why you felt the intervention would be successful in aiding you to stop smoking?

Do you feel the intervention will be successful in helping other staff stop smoking?

Can you tell me why you feel the intervention will/will not help other staff stop smoking?

Can you tell me how did the intervention improve your knowledge of smoking, and aid in you stop smoking?

**Self-Efficacy**: *the participants' confidence that they can perform the behaviour(s) required to participate in the intervention*

What skills and level of confidence do you think is needed to be successful in stop smoking?

Can you tell me if you believe you have those skills and confidence?

Why is that? Example?

Looking back, did you have any uncertainties around your ability to engage with the service?

Would you recommend the service to another staff member?

Were there any unexpected actions from this intervention that you have not expected?

Can you explain?

Would you feel confident approaching the service if you required more support?

Do you feel there was a sufficient amount of NRT/vape offered?
