## Supplementary Materials 2 - Survey for "The Acceptability of a Tobacco Dependency Treatment for NHS Staff in the North East of England: A Mixed-Methods Study"

NIHR | Applied Research Collaboration North East and North Cumbria

NHS North East and North Cumbria

**Evaluating the North East and North Cumbria NHS Staff Tobacco Dependency Offer**

Survey

Gender

| Male | Female | Non-binary/third gender | Prefer not to say |
|---|---|---|---|

Ethnicity

| White – British | White – Irish | White – any other white background | Mixed – White and Black Caribbean | Mixed – White and Black African | Mixed – White and Asian |
|---|---|---|---|---|---|

| Mixed – any other mixed background | Asian or Asian British – Indian | Asian or Asian British – Pakistani | Asian or Asian British – Bangladeshi | Asian or Asian British – any other Asian background | Black or Black British - Caribbean |
|---|---|---|---|---|---|

| Black or Black British – African | Black or Black British – any other Black background | Other Ethnic Groups – Chinese | Other Ethnic Groups – any other ethnic group |
|---|---|---|---|

If other, please state

Age

What is the first half of your home postcode? (i.e., NE30)

Which local authority did you access?

| Durham County Council | Gateshead Council (Including QEF | Hartlepool Council (provided by | Newcastle Council (provided by CGL) | North Cumbria (provided by Gateshead |
|---|---|---|---|---|

| (provided by ABL Health) | outpatient pharmacy) | Hartlepool Support Hub) | | Health Staff Team or NHS Smoke Free App) |
|---|---|---|---|---|

| North Tyneside Council | Northumberland Council | South Tees Stop Smoking Service | South Tyneside Council | Stockton-on-Tees Council |
|---|---|---|---|---|

| Sunderland Council | Smokefree App | Smokefree Staff Team – Gateshead Health | I don't know |
|---|---|---|---|

Work setting

| Allied Healthcare Professionals | Nursing and Midwifery Registered | Community Services | Clinical Support Staff | Admin and Clerical | Porting and Estates |
|---|---|---|---|---|---|

| Domestic Services and Catering | Corporate Services | Directors and Senior Management | Medical Professional | Other |
|---|---|---|---|---|

If other, please state

1. Affective attitude – *how an individual feels about the intervention*

How comfortable did you feel engaging with the NHS Staff Tobacco Dependency Offer (STDO)?

| Very uncomfortable | Uncomfortable | No opinion | Comfortable | Very comfortable |
|---|---|---|---|---|
| 1 | 2 | 3 | 4 | 5 |

2. Burden – *the amount of effort required to participate in the intervention*

How much effort did it take to engage with the NHS Staff Tobacco Dependency Offer (STDO)?

| No effort at all | A little effort | No opinion | A lot of effort | Huge effort |
|---|---|---|---|---|
| 1 | 2 | 3 | 4 | 5 |

3. Ethicality – *the extent to which the intervention has good fit with an individual's value system.*

There are moral or ethical consequences to engage with the NHS Staff Tobacco Dependency Offer (STDO)?

| Strongly disagree | Disagree | No opinion | Agree | Strongly agree |
|---|---|---|---|---|
| 1 | 2 | 3 | 4 | 5 |

4. Perceived effectiveness – *the extent to which the intervention is perceived to have achieved its objective*

The NHS Staff Tobacco Dependency Offer (STDO) has aided me in a quit attempt

| Strongly disagree | Disagree | No opinion | Agree | Strongly agree |
|---|---|---|---|---|
| 1 | 2 | 3 | 4 | 5 |

5. Intervention coherence – *the extent which the participant understands how the intervention works*

It is clear to me how the NHS Staff Tobacco Dependency Offer (STDO) will help me in a quit attempt

| Strongly disagree | Disagree | No opinion | Agree | Strongly agree |
|---|---|---|---|---|
| 1 | 2 | 3 | 4 | 5 |

6. Self-efficacy – *a participants confidence that they can perform behaviours required to participate in the intervention*

How confident did you feel about engaging with the NHS Staff Tobacco Dependency Offer (STDO)?

| Very unconfident | Unconfident | No opinion | Confident | Very confident |
|---|---|---|---|---|
| 1 | 2 | 3 | 4 | 5 |

7. Opportunity costs – *the benefits, profits or values that would have to be given up to engage with the intervention*

Engaging with the NHS Staff Tobacco Dependency Offer (STDO) interfered with my other priorities

| Strongly disagree | Disagree | No opinion | Agree | Strongly agree |
|---|---|---|---|---|
| 1 | 2 | 3 | 4 | 5 |

8. General acceptability

How acceptable was the NHS Staff Tobacco Dependency Offer (STDO) to you?

| Completely unacceptable | Unacceptable | No opinion | Acceptable | Completely acceptable |
|---|---|---|---|---|
| 1 | 2 | 3 | 4 | 5 |

We will be conducting a small number of informal interviews to help us further explore and understand the views and experiences of NHS staff who have accessed the staff tobacco dependency offer. As a thank you for your time, a £15 Love2Shop voucher will be made available to you upon completion of the interview. If you are happy to and would like to receive further information about this, please include your details below.

Name

Email

Telephone
